# Evaluation of the psychometric properties of the Episodic Disability Questionnaire (EDQ) among women living with HIV in the United Kingdom: a self-reported repeated measure study

**DOI:** 10.1101/2025.10.30.25339187

**Authors:** DA Brown Obe, S Tariq, M Boffito, D Asboe, A Milinkovic, N Nwokolo, C Flavell, S Strachan, L Avery, KK O’Brien, R Harding

**Affiliations:** Chelsea and Westminster Hospital NHS Foundation Trust, London, United Kingdom; University College London, London, United Kingdom; Central and North West London NHS Foundation Trust, London, United Kingdom; James Cook University, Townsville, Queensland, Australia; Sophia Forum, London, United Kingdom; University Health Network, Toronto, Canada; Dalla Lana School of Public Health, University of Toronto, Toronto, Canada; Department of Physical Therapy, Temerty Faculty of Medicine, University of Toronto, Toronto, Canada; Institute of Health Policy, Management and Evaluation, Dalla Lana School of Public Health, University of Toronto, Toronto, Canada; Rehabilitation Sciences Institute, University of Toronto, Toronto, Canada; Cicely Saunders Institute, Florence Nightingale Faculty of Nursing Midwifery and Palliative Care, King’s College London, London, United Kingdom

**Keywords:** HIV, women, disability, psychometric properties, patient-reported outcomes

## Abstract

**Background:** Disability is an increasingly common health outcome as women age with HIV and multimorbidity. The Episodic Disability Questionnaire (EDQ) measures the presence, severity, and episodic nature of disability across six domains. We evaluated EDQ properties among women living with HIV in the United Kingdom.

**Methods:** Participants in the Positive Transitions Through the Menopause (PRIME) study completed the EDQ at two timepoints (1 week apart), criterion measures (WHODAS 2.0, EQ-5D-5L, Work and Social Adjustment Scale), and a demographic questionnaire. We evaluated internal consistency (Cronbach alpha ≥0.7), test-retest reliability (ICC >0.7), measurement precision (Minimum Detectable Change (MDC) 95%), and construct validity (≥75% a priori hypotheses met). Disability prevalence was assessed using WHODAS 2.0 (moderate threshold) and Equality Act Disability Definition (severe threshold).

**Results:** Of 104 participants (median age 56 years, 65% Black ethnicity), 93 (89%) completed the EDQ twice. Median duration since HIV diagnosis was 23 years; 98% had undetectable viral loads and 86% reported multimorbidity. Cronbach’s alpha ranged from 0.83 (social domain) to 0.92 (daily activities domain). ICC ranged from 0.70 (physical domain) to 0.91 (daily activities domain). Precision was highest in daily activities domain (MDC95%: 6.10) and lowest in mental-emotional domain (MDC95%: 11.52). Eighty percent (n=47/59) of construct validity hypotheses were met. Disability prevalence was 79.81% (95%CI 70.57, 86.79) moderate and 41.75% (32.24, 51.88) severe disability.

**Conclusions:** The EDQ possesses internal consistency, test-retest reliability, and construct validity with varied precision among women living with HIV. Results can support research and clinical practice to measure disability and evaluate interventions.

## INTRODUCTION

With access to effective antiretroviral therapy, people living with HIV are increasingly reaching older age in the United Kingdom (UK) and are more likely to experience multimorbidity and disability [1-3], presenting significant health-related challenges. Disability is defined by adults living with HIV as any physical, cognitive, mental-emotional symptoms, difficulties with day-to-day activities, challenges to social inclusion, and uncertainty about future health, that may persist or fluctuate on a daily basis or over the longer course of living with HIV [4, 5].

Functioning and disability are important for monitoring the performance of health strategies in health systems [6]. Measuring disability is additionally important for determining the impact of health challenges, improving communication between providers and patients, and evaluating interventions [7-9].

The Episodic Disability Questionnaire (EDQ) is a generic 35-item patient-reported outcome measure of disability [10] based on a conceptual framework of episodic disability [4, 11]. It is derived from the HIV-specific Short-Form HIV Disability Questionnaire (SF-HDQ) [12] and HIV Disability Questionnaire (HDQ) [13], which both possess validity, reliability, and sensibility for use among adults living with HIV in Canada, Ireland, the United States (US) and the UK [14-18]. The EDQ has demonstrated internal consistency, construct validity, and test–retest reliability, with limited precision when administered electronically across a sample comprised of primarily men (83%) living with HIV [10].

In the UK, people with disabilities experience inequities in education, employment, living standards, housing, well-being, loneliness, and are more commonly victims of crime than the general population [19]. Women are more likely to experience disability than men [19-21] and women living with HIV in the UK experience higher disability severity compared to men [3]. Considering the gendered dimensions of disability and the under-representation of women in HIV research [22], it is essential to evaluate the psychometric properties of disability measurement tools among women. We aimed to assess the EDQ for its ability to measure disability experienced by women living with HIV in the UK, specifically internal consistency, test-retest reliability, precision of measurement, and construct validity. We also aimed to measure disability prevalence and report disability profiles among women living with HIV.

## METHODS

We conducted a cross-sectional repeated measurement study involving the administration of the EDQ and criterion measures with women living with HIV in England, UK. We followed the COnsensus-based Standards for the selection of health Measurement Instruments (COSMIN) guidelines for assessment and reporting of psychometric properties of the EDQ [23].

### Study setting

This study was conducted at ten NHS outpatient HIV clinical settings in three cities in England, UK. We received ethics approval from the London City & East Research Ethics Committee (REC reference: 22/PR/1483) and Health Research Authority (IRAS: 318781). This study was included in the NIHR CRN Portfolio (CPMS ID: 54554).

### Participants

We recruited women living with HIV who had previously participated in the Positive Transitions Through the Menopause (PRIME) study, regardless of menopausal status [24]. Potential participants were eligible if they had participated in PRIME, and were therefore living with HIV, female sex, aged 45-60 years on entry to PRIME, and had provided consent to be contacted about future research. Exclusion criteria were the inability to give consent or complete questionnaires in English. Participants were recruited via their local clinical care team. Written informed consent was obtained from participants at the initial study information and consent page of the questionnaire administration.

### Data collection

Between March 2023 to January 2024 we electronically administered the EDQ followed by three criterion measures (World Health Organization Disability Assessment Schedule (WHODAS 2.0) [25], EQ-5D-5L [26], and Work and Social Adjustment Scale (WSAS) [27]) and a demographic questionnaire using Qualtrics software (https://www.qualtrics.com). Participants completed the questionnaires in-person via a computer or tablet at the recruiting clinical site, or remotely via a link in an email or Short Message Service (SMS) text. One week later, we emailed participants with a link requesting them to complete the EDQ only. At this time, we asked whether participants had a major change in their health status since their last EDQ completion and if yes, to describe this change. Study authors did not have access to information that could identify individual participants during or after data collection.

### Questionnaires

#### Episodic Disability Questionnaire

The EDQ is a generic patient-reported outcome measure (PROM) comprising 35-items spanning six domains: i) physical (10 items); ii) cognitive (3 items); iii) mental-emotional (5 items), iv) uncertainty about future health (5 items), v) difficulties carrying out day-to-day activities (5 items), and vi) challenges to social inclusion (7 items) [10]. For each item, individuals indicate the extent they are experiencing a specific health-related challenge on the day of assessment using item-specific severity scales: 0-4 (32 items), 0-3 (1 item), and 0-2 (2 items). They also report whether each challenge fluctuated over the past week with a binary episodic score (yes/no). The presence score is derived by dichotomising severity as present (severity 1–4) or absent (severity of 0). Severity and presence domain scores are calculated using the Rasch analysis algorithm (score range: 0-100) [12]. Higher scores indicate greater presence, severity and episodic nature of disability.

#### World Health Organization Disability Assessment Schedule

The WHODAS 2.0 is a 12-item self-administered generic questionnaire of functioning and disability applicable across cultures in adult populations, and directly linked to the International Classification of Functioning, Disability and Health (ICF) [25, 28]. The WHODAS 2.0 assesses difficulty in performing specific functions over the previous 30 days across six disability domains: i) cognition, ii) mobility, iii) self-care, iv) getting along, v) life activities and vi) participation. Individuals answer on a 5-point ordinal scale (range 0–4) with higher scores indicating increasing difficulty completing the task. In simple scoring, scores are summed to provide an answer out of 48, with higher scores suggesting greater disability [25]. Complex scoring (or item response theory-based scoring) weights individual item severity, providing a disability range from 0 (no disability) to 100 (total disability) [25]. The WHODAS possesses internal consistency, test–retest reliability, validity and cross-cultural applicability [28, 29]. The WHODAS is validated in people with chronic diseases [30] and people living with HIV [31]. Categorisation thresholds have been developed to identify people living with HIV experiencing disability (score ≥2) [3, 32, 33], and any level of functional limitation (score ≥1) [34, 35]. In the UK, scores ≥2 have been defined as “moderate” disability among people living with HIV [3].

#### EQ-5D-5L

The EuroQOL five dimensions five-level questionnaire is a generic self-reported measure of health-related quality of life, comprising five dimensions: i) mobility; ii) self-care; iii) usual activities; iv) pain/discomfort; and v) anxiety/depression [26, 36]. Each dimension has 5 levels: no problems, slight problems, moderate problems, severe problems, and extreme problems. The digits for the five dimensions can be combined into a 5-digit number that describes the patient’s health state, or represented by a single summary number known as the index value, with a range approximately −0.285 to 1.000 [26, 37]. The EQ-5D-5L is an extensively used generic measure of health-related quality of life in HIV research and across different diseases worldwide [38, 39]. Psychometric data support its use [40], and it has been used within national HIV reporting and the PRIME study [24, 41].

#### Work and Social Adjustment Scale

The WSAS is a generic measure of perceived impairment in work or social functioning resulting from a health problem [27]. The 5 items are scored on a ordinal scale from 0 (not at all) to 8 (very severe), to identify challenges in domains: i) work; ii) home management; iii) social leisure activities; iv) private leisure activities; and v) relationships with others. Scores range from 0 to 40, with lower scores indicating better adjustment. WSAS scores above 20 suggest severe functional impairment, scores between 10 and 20 suggest moderately severe and scores below 10 suggest mild [27, 42]. The WSAS is a unidimensional scale, suggesting the scores cannot be compared across groups of different health conditions, with overall high internal consistency among people living with HIV [42] and used to assess functional impairment among people living with HIV and painful peripheral neuropathy [43].

#### Demographic Questionnaire

The demographic questionnaire included 35 items comprised of demographic (eg: age, sex, gender, ethnicity), health (eg: time since HIV diagnosis, viral load, concurrent health conditions, menopause, and general health status), and social characteristics (eg: living arrangements, work status, social security or benefits), and the UK Equality Act Disability Definition (EADD) questions [44, 45]. The EADD reflects legal definitions in the Equality Act 2010 and defines a person as disabled if they have a physical or mental impairment and that impairment has a substantial and long-term adverse effect on their ability to carry out day-to-day activities [46]. The classification questions making up the EADD are: (a) “*do you have any physical or mental health conditions or illnesses lasting or expecting to last 12-months or more?*”; (b) “*do any of your conditions of illnesses reduce your ability to carry out day-to-day activities?*”. A person is counted as disabled if they answer “*yes*” to both classification questions. The EADD has been used to define “severe” disability among people living with HIV in the UK [3].

### Analysis

We calculated median (interquartile ranges (IQR)) EDQ scores. Severity and presence domain scores were calculated using the Rasch analysis algorithm (score range: 0-100) [12]. Episodic domain scores included a simple sum transformed on a scale 0-100. Higher scores indicated greater presence, severity and episodic nature of disability. We calculated median WHODAS 2.0 domain scores, EQ-5D-5L scores and WSAS scores as per guidelines. For the demographic questionnaire, we calculated descriptive statistics including frequencies (%) for categorical variables and median and IQR for continuous variables. Analysis was conducted using R [@R-psych] version 4.4.0.

#### Internal Consistency

We calculated Cronbach’s alpha (severity domain) and Kuder-Richardson-20 (KR-20) (presence and episodic domains) for time 1 (T1) with 95% confidence intervals (CI) (≥0.7 acceptable) [47].

#### Test-Retest Reliability

We calculated Intra Class Correlations (ICCs) with 95% CI using T1 and time 2 (T2) EDQ scores based on Shrout and Fleiss’ ICC2 (absolute agreement with random raters) (lower bound CI of > 0.7 acceptable) [47]. We calculated ICCs for the entire sample indicating no change in their health status. Our test–retest assessment focused on EDQ presence and severity scales, as the episodic scale refers to fluctuations in disability in the past week, and we did not expect consistency in this scale.

#### Measurement Precision

We calculated the Standardised Error of Measurement (SEM) using Wywrich criteria [48], 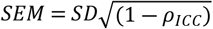 where *ρ*_*ICC*_ is the test-retest reliability. The Minimum Detectable Change (MDC) was calculated for 95% CI (MDC95%) using the method proposed by Haley [49], 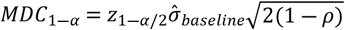 where ρ is the test-retest reliability ICC, 1 - α is the level of confidence, and 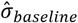 is the standard deviation of the measure at T1.

#### Construct Validity

We examined correlations for 59 total *a priori* hypotheses: 6 primary and 53 secondary. Primary hypothesis theorised relationships between EDQ domains’ severity scores and WHODAS 2.0 total scores. Secondary hypothesis theorised relationships between EDQ and WHODAS, EQ-5D-5L, and WSAS criterion measure sub-scales, and known groups of participants completing EDQ “on a good day” and with ≥ 2 concurrent health conditions. We derived *a priori* hypotheses from earlier construct validity assessments of the EDQ [10]. Spearman correlation coefficients of |≥ 0.30|, |≥ 0.50| and |≥ 0.70| were defined as ‘weak’, ‘moderate’ and ‘strong’, respectively [50]. If the lower bound of the confidence interval for the Spearman correlation was greater than the pre-specified level, the criteria were considered to be met. Wilcoxon rank sum test for a test of difference in medians between the known groups was significant at a level of <0.05, for the criteria to be considered met. Construct validity was defined as ≥75% confirmed hypotheses [50].

#### Disability Prevalence

We estimated disability prevalence frequencies (%) and 95% CI, representing a range of severity [51]. The threshold for experiencing “moderate” disability was defined as WHODAS scores ≥2, representing at least two mild/moderate or one moderate/severe limitation on WHODAS items [3, 32-35]. The threshold for experiencing “severe” disability was defined as self-rating “yes” to both EADD classification questions [3].

#### Disability Profile

We calculated the frequency (%) for WHODAS simple scores in response to all 12 items, presence of any functional limitation (score ≥1) per WHODAS domain, WHODAS total number of limitations, and WHODAS difficulty levels. For WHODAS simple and complex sum scores, we calculated mean (standard deviation (SD)) and median (IQR as 25^th^-75^th^ percentile) to align with normative data and existing literature [3, 52, 53]. We calculated median, lower quartile (LQ) and upper quartile (UQ), and range for EDQ presence, severity, and episodic scores per EDQ domain.

#### Sample Size

To detect a weak correlation |r=0.30| between EDQ and criterion scores, with a power of 0.80 and alpha of 0.05 required 85 participants [54]. To account for questionnaires with missing responses, loss to follow-up at T2, and recruiting 25% eligible participants from each site, our targeted sample size was 104 women living with HIV.

## RESULTS

One hundred and four participants completed the questionnaires at T1, of which 93 (89%) completed the EDQ at T2, a median 11 days (IQR 8-19) after T1. Of the 93 completing T2, 59 (63%) participants reported no change in health status and were included in the test-retest reliability assessment. Among the 59 included, 51 (86%) reported completing T1 and T2 questionnaires on a good day, and 8 (14%) reported a bad day for both T1 and T2.

### Participant Characteristics

See Table 1 for characteristics of participants. All participants were female sex assigned at birth, median age 56 years (IQR 54-58), with a median duration of 23 years since HIV diagnosis (IQR 18-27). Most participants were of Black, Black British, Caribbean or African ethnicity n=67 (65%), taking anti-retroviral therapy n=102 (99%), had an undetectable viral load <200 copies/ml n=88 (98%), and were living with a median of 5 comorbidities (IQR 3-8), representing multi-morbidity (≥2 co-morbidities) n=89 (86%).

**Table 1:**
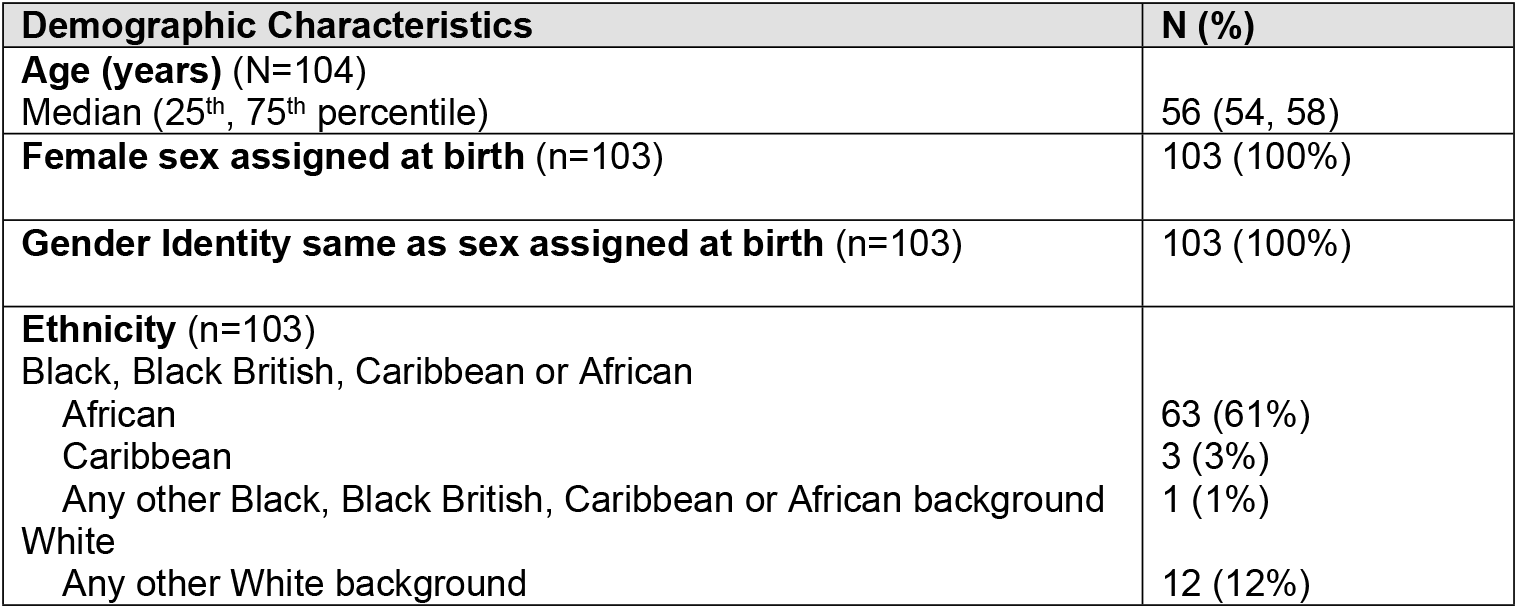
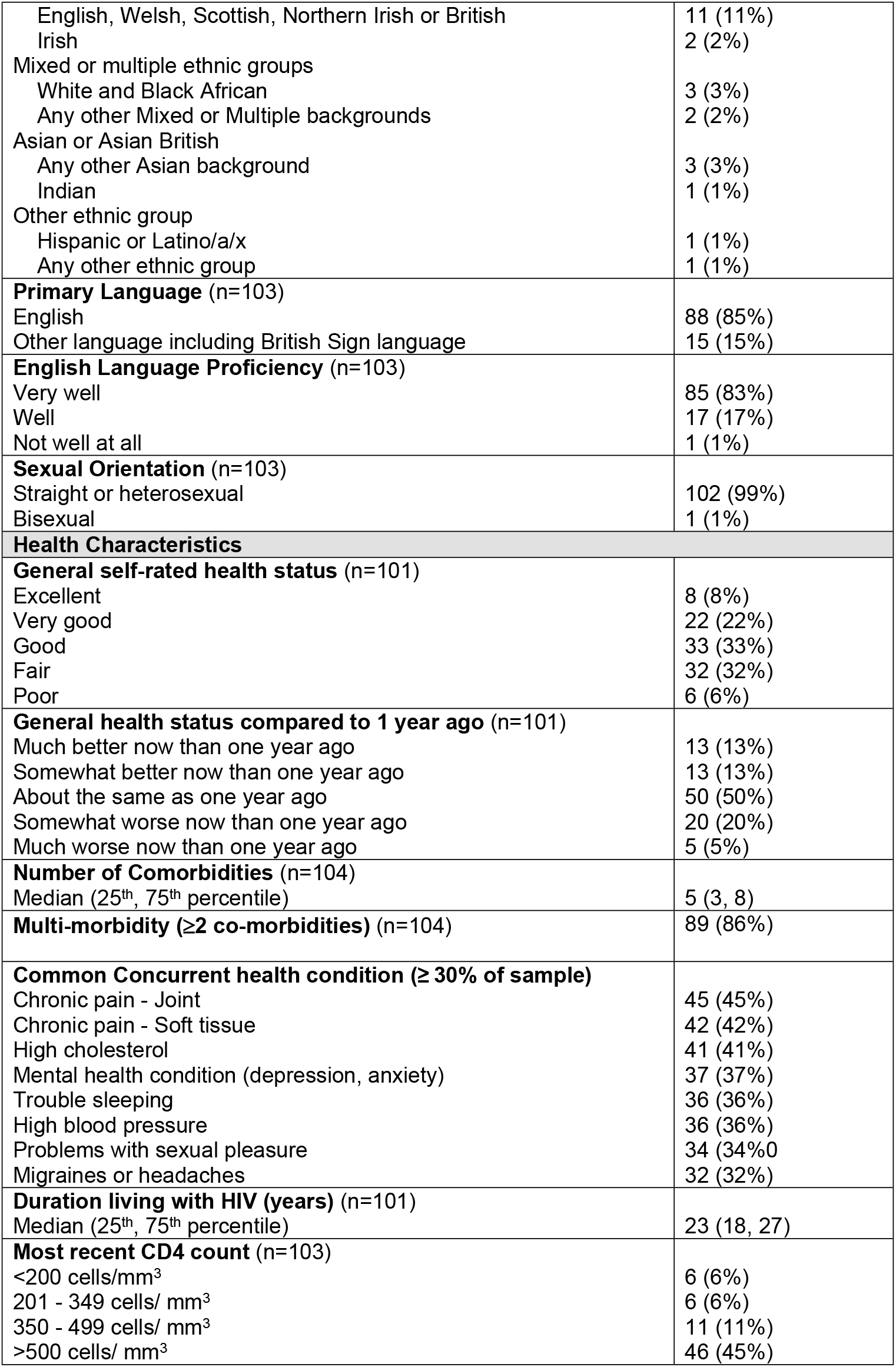
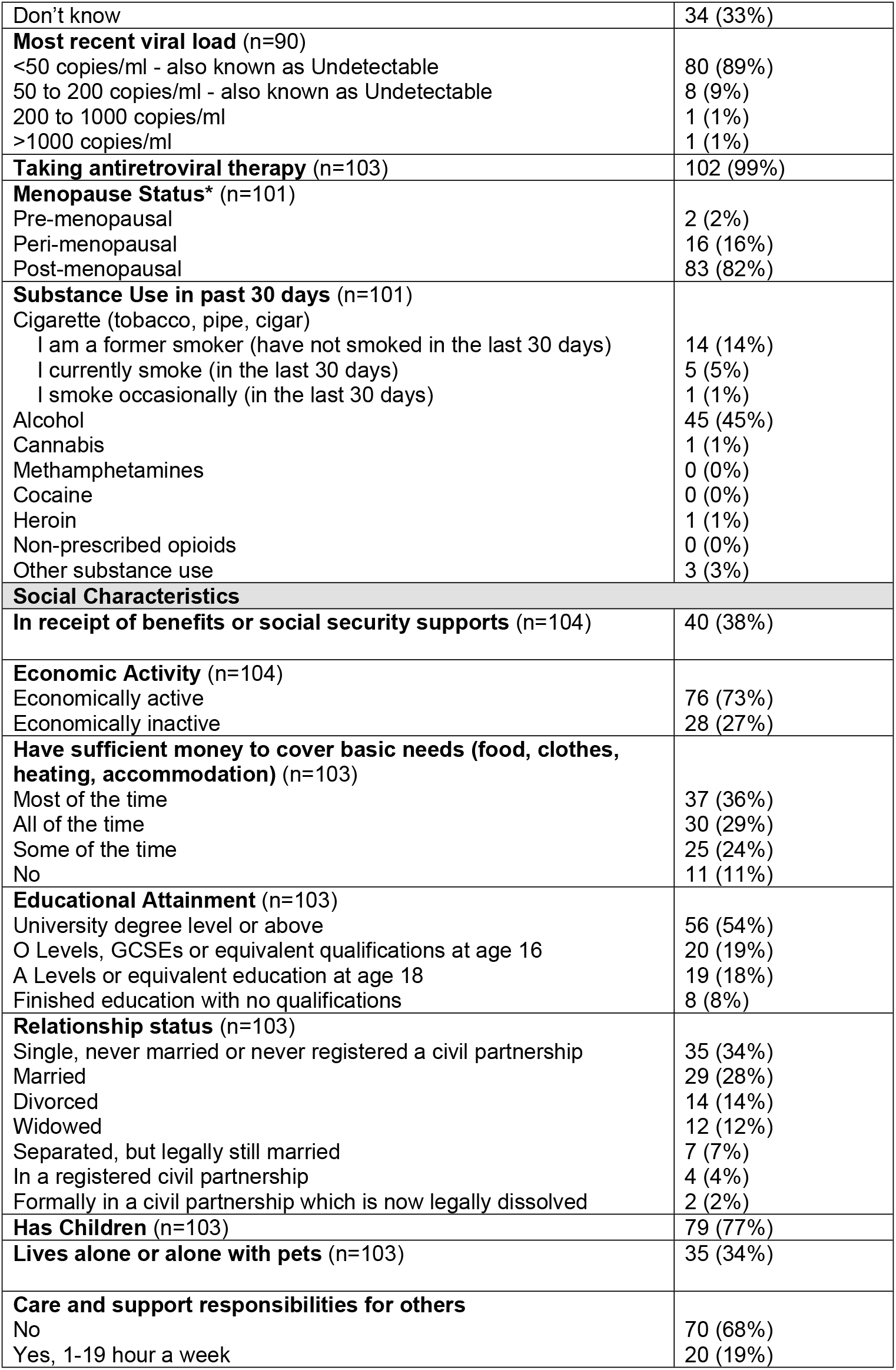
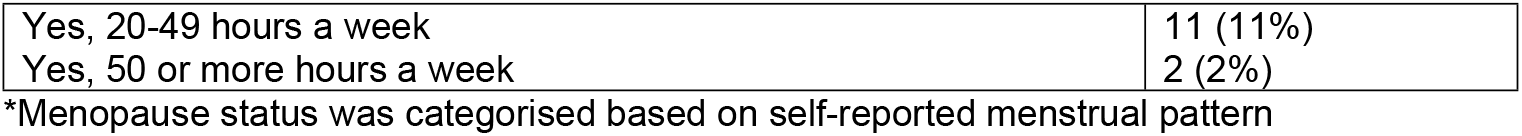
Participants’ demographic, health, and social characteristics.

### Criterion Measures

WHODAS “simple” sum scores were mean 12.6 out of 48 (SD; 11.9), median 9.0 (IQR; 2.0-22.2). WHODAS “complex” sum scores were mean 26.3 out of 100 (SD; 24.8), median 18.8 (IQR; 4.2-46.3). The EQ-5D-5L median index scores were 0.7 (IQR: 0.5-0.8). See Supplementary File Table S.1 for frequencies (%) for each ED-5D-5L domain/item. The WSAS scores were mean 9.9 out of 40 (SD;11.8), median 5 (IQR: 0-16).

### Internal Consistency

The EDQ met criteria for internal consistency across domains in the presence, severity and episodic scales (ICC ≥0.7). Cronbach’s alpha for EDQ severity scores range from 0.83 (social domain) to 0.92 (daily activities domain), for EDQ presence scores ranged from 0.75 (uncertainty domain) to 0.90 (daily activities domain), and for EDQ episodic scores ranged from 0.70 (social domain) to 0.81 (daily activities domain) (Table 2). Lower bound CIs were ≥0.7, except for the uncertainty domain (0.67) of the EDQ presence scale and social domain (0.61) of the EDQ episodic scale (Table 2).

**Table 2:** Internal consistency for Episodic Disability Questionnaire (EDQ) domain scores (n=104 participants)

| EDQ Domain | EDQ Severity<br>Cronbach's alpha<br>(95% CI) | EDQ Presence<br>KR-20<br>(95% CI) | EDQ Episodic<br>KR-20<br>(95% CI) |
| --- | --- | --- | --- |
| Physical | 0.87 (0.83, 0.91) | 0.80 (0.74, 0.85) | 0.78 (0.71, 0.84) |
| Cognitive | 0.89 (0.85, 0.92) | 0.81 (0.74, 0.87) | 0.79 (0.71, 0.85) |
| Mental-emotional | 0.91 (0.88, 0.93) | 0.82 (0.76, 0.87) | 0.80 (0.73, 0.85) |
| Uncertainty | 0.86 (0.81, 0.90) | 0.75 (0.67, 0.82) | 0.80 (0.73, 0.85) |
| Daily activities | 0.92 (0.89, 0.94) | 0.90 (0.86, 0.93) | 0.81 (0.74, 0.86) |
| Social inclusion | 0.83 (0.77, 0.87) | 0.79 (0.73, 0.85) | 0.70 (0.61, 0.78) |

### Test-Retest Reliability

Overall, the EDQ met criteria for test-retest reliability for EDQ severity domains with ICCs ranging from 0.70 (physical domain) to 0.91 (daily activities domain) and for EDQ presence domains ranging from 0.72 (physical domain) to 0.89 (daily activities domain) (Table 3). Lower bound CIs were >0.7, except for the physical domain (0.53) of the EDQ severity scale and physical (0.57), cognitive (0.60) and uncertainty (0.63) domains of the EDQ presence scale (Table 3).

**Table 3:** Test-retest reliability for the Episodic Disability Questionnaire (EDQ) domains for severity and presence scales (n=59)

| EDQ Domain | EDQ Severity<br>ICC (95% CI) | EDQ Presence<br>ICC (95% CI) | EDQ Episodic*<br>ICC (95% CI) |
| --- | --- | --- | --- |
| Physical | 0.70 (0.53, 0.81) | 0.72 (0.57, 0.82) | 0.69 (0.47, 0.82) |
| Cognitive | 0.81 (0.70, 0.88) | 0.74 (0.60, 0.84) | 0.36 (0.12, 0.56) |
| Mental-emotional | 0.78 (0.65, 0.86) | 0.84 (0.75, 0.90) | 0.49 (0.27, 0.66) |
| Uncertainty | 0.89 (0.82, 0.94) | 0.76 (0.63, 0.85) | 0.31 (0.07, 0.52) |
| Daily activities | 0.91 (0.86, 0.95) | 0.89 (0.82, 0.93) | 0.20 (-0.05, 0.43) |
| Social inclusion | 0.87 (0.78, 0.92) | 0.82 (0.72, 0.89) | 0.25 (-0.01, 0.47) |

\* For information only as consistency is not expected in the episodic scale **Measurement**

### Precision

The EDQ severity scale for each domain demonstrated the highest precision, whereby the MDC95% ranged from 6.10 (daily activities domain) to 11.52 (mental-emotional domain), followed by the presence scale, whereby MDC95% ranged from 12.84 (social inclusion domain) to 21.32 (cognitive domain). The episodic scale MDC95% ranged from 13.75 (social inclusion domain) to 27.58 (cognitive domain) (Table 4).

**Table 4:** Minimum Detectable Change (MDC) for Episodic Disability Questionnaire (EDQ) scales (n=104 participants)

| EDQ Domain | EDQ Severity<br>SEM (MDC95%) | EDQ Presence<br>SEM (MDC95%) | EDQ Episodic<br>SEM (MDC95%) |
| --- | --- | --- | --- |
| Physical | 9.31 (18.24) | 13.25 (25.98) | 14.00 (27.45) |
| Cognitive | 10.38 (20.34) | 21.32 (41.79) | 27.58 (54.06) |
| Mental-emotional | 11.52 (22.59) | 14.27 (27.98) | 21.93 (42.97) |
| Uncertainty | 6.92 (13.56) | 14.55 (28.52) | 22.55 (44.21) |
| Daily activities | 6.10 (11.96) | 13.71 (26.87) | 23.24 (45.56) |
| Social inclusion | 6.75 (13.24) | 12.84 (25.16) | 13.75 (26.95) |

### Construct Validity

Eighty percent (47/59) of all hypotheses were met, including all six primary hypotheses (100%) and 41/53 (77%) secondary hypotheses confirmed, supporting construct validity for use with women living with HIV in the UK [Supplementary File Table S.2].

### Disability Prevalence

The estimated prevalence of moderate disability was 79.81% (95%CI: 70.57, 86.79) with n=83 scoring ≥2 on WHODAS. The estimated prevalence of severe disability was 41.75% (95%CI: 32.24, 51.88) with n=43 self-rating “yes” to both EADD classification questions.

### Disability Profile

The WHODAS simple scores in response to all 12 items are shown in Supplementary File Table S.3. Frequency of any functional limitation (score ≥1) within each of the six WHODAS disability domains were; challenges to social participation (n=129, 62%), mobility challenges (n=122, 59%), challenges to life activities (n=116, 56%), cognitive health challenges (n=103, 50%), challenges getting along (n=90, 43%), and challenges with self-care (n=72, 35%). Any level of functional limitation (WHODAS score ≥1) was reported by n=90 (87%), whereby n = 9 (9%) scored one limitation, n=10 (10%) two limitations, n=7 (7%) three limitations, n=64 (62%) four or more limitations, and n=19 (18%) scoring all twelve limitations. Difficulty levels across all WHODAS items were “no difficulty’ n=616 (49%), “mild difficulty” n=203 (16%), “moderate difficulty” n=247 (20%), “severe difficulty” n=114 (9%), and “extreme difficulty/cannot do” n=68 (5%). The T1 EDQ domain scores are shown in Table 5. The most severe, present, and episodic EDQ disability domains at T1 were “uncertainty”, “uncertainty”, and “physical symptoms and impairments” respectively, with n=84 (81%) reported completing the EDQ on a good day at T1.

**Table 5:**
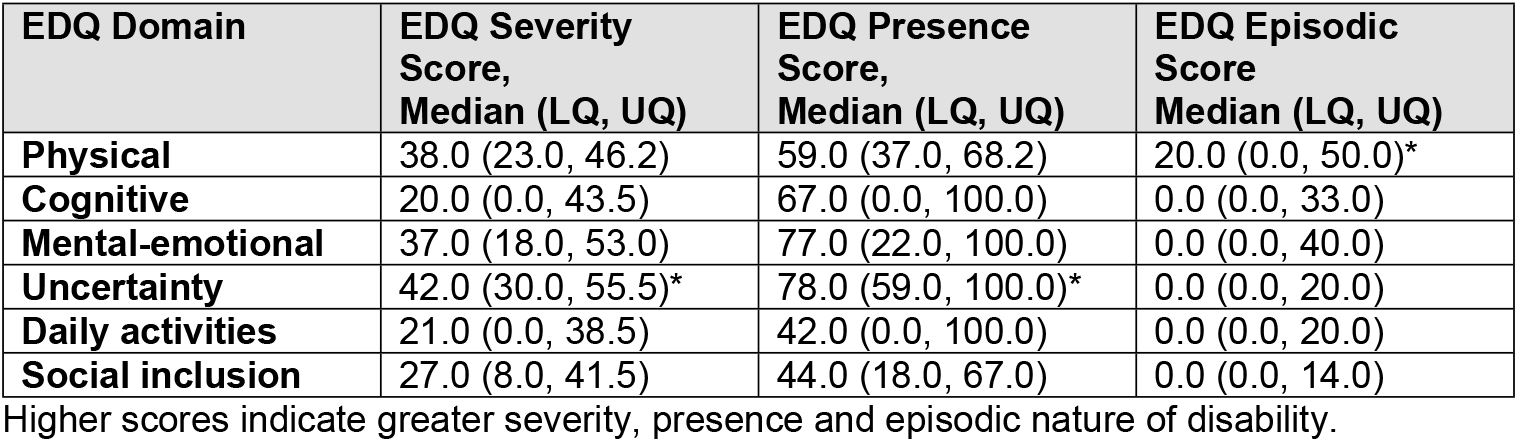
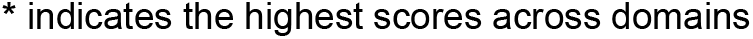
Median Time 1 EDQ Domain Scores (n=104 participants)

| EDQ Domain | EDQ Severity<br>Score,<br>Median (LQ, UQ) | EDQ Presence<br>Score,<br>Median (LQ, UQ) | EDQ Episodic<br>Score<br>Median (LQ, UQ) |
| --- | --- | --- | --- |
| Physical | 38.0 (23.0, 46.2) | 59.0 (37.0, 68.2) | 20.0 (0.0, 50.0)* |
| Cognitive | 20.0 (0.0, 43.5) | 67.0 (0.0, 100.0) | 0.0 (0.0, 33.0) |
| Mental-emotional | 37.0 (18.0, 53.0) | 77.0 (22.0, 100.0) | 0.0 (0.0, 40.0) |
| Uncertainty | 42.0 (30.0, 55.5)* | 78.0 (59.0, 100.0)* | 0.0 (0.0, 20.0) |
| Daily activities | 21.0 (0.0, 38.5) | 42.0 (0.0, 100.0) | 0.0 (0.0, 20.0) |
| Social inclusion | 27.0 (8.0, 41.5) | 44.0 (18.0, 67.0) | 0.0 (0.0, 14.0) |

Higher scores indicate greater severity, presence and episodic nature of disability.
\* indicates the highest scores across domains

## DISCUSSION

The EDQ possesses internal consistency, test-retest reliability, and construct validity with varied precision among women living with HIV in the UK. This work goes beyond past disability measurement property assessments, which primarily included samples of men living with HIV [10, 12, 14-18]. These results are the first known to focus on the EDQ measurement properties with women.

The EDQ presence, severity and episodic scales possessed internal consistency with all lower bound CIs of Cronbach’s alphas >0.70, except the uncertainty domain in the EDQ presence scale (0.67) and social domain in the EDQ episodic scale (0.61), suggesting the domains are homogenous and collectively measure the broader construct of disability in women living with HIV in the UK. This aligns with internal consistency assessment of the EDQ in Canada, Ireland, the UK and the US, whereby Cronbach’s alphas ranged from 0.72-0.91 [10].

Construct validity was supported, with 80% of overall *a priori* hypotheses fulfilled, including all primary and 77% secondary outcomes. This level of hypothesis confirmation, alongside a conservative approach to evaluating both primary and secondary criteria, reinforces the EDQ’s capacity to measure disability in alignment with previous literature [10, 16]. Considering gendered dimensions of disability, these findings provide clinicians and researchers assurance of the EDQ’s ability to measure disability experienced by women living with HIV in the UK.

Our assessment of test-retest reliability of the EDQ severity and presence scales demonstrated ICCs >0.70. Lower bound CIs for all ICCs were >0.70, except the physical (0.53) domain of the EDQ severity scale and physical (0.57), cognitive (0.60) and uncertainty (0.63) domains of the EDQ presence scale. This assessment, involving only two points in time, highlights the potential for daily fluctuations in disability, which may affect T2 EDQ score interpretations and potentially underestimate the EDQ’s test-retest reliability. Further longitudinal assessments with additional repeated measures over time could enhance the administration of this tool throughout routine HIV clinical care visits. It has been suggested that the EDQ may be well-suited for group-based or program evaluation purposes [10]. However, it is ultimately for clinicians to determine the acceptable level of error in clinical practice, depending on the intended purpose for EDQ administration (e.g. service referrals, eligibility for disability benefits [14, 55]).

The EDQ demonstrated higher precision within the severity domain among women living with HIV, underscoring its potential utility in accurately interpreting longitudinal changes in disability. The severity scale offers clinicians and researchers a reference point for discerning minimal yet significant changes in disability over time, surpassing day-to-day variability and measurement error.

Our disability prevalence estimates among this sample of working-age women living with HIV in the UK demonstrate that 4 in 5 (79.81%) women experienced moderate disability, and approximately 2 in 5 (41.75%) experienced severe disability. The severe disability threshold (EADD) follows the core definition of disability in the Equality Act 2010, permitting comparison to the general population. Our disability estimates exceed proportions of disability among women and working-age adults in the UK general population, trending towards proportions among state-pension age adults [56]. They also exceed disability prevalence among a sample comprised primarily of men living with HIV in the UK [3]. Notably, 1 in 6 (18%) participants reported via the EADD that their conditions or illnesses limited their ability to carry out day-to-day activities, despite indicating no physical or mental health conditions expected to last 12-months or more. Given that all participants were living with HIV, this may represent underreporting of the severe disability prevalence and highlights potential limitations of duration-based criteria in capturing episodic or fluctuating disability experiences. This sample of women also had more concurrent health conditions (median 5) compared to previous estimates among men (median 2) [3], underscoring the gendered complexity of ageing with HIV and multimorbidity. These findings reinforce the importance of using disability measures that account for episodic health challenges, particularly among populations ageing with HIV.

Our findings demonstrate that women living with HIV experience multidimensional and episodic disability in the context of antiretroviral therapy and viral suppression. This study also provides the first known disability profiles of women living with HIV in the UK, whereby the most severe and present disability domain was uncertainty or worry about future health. This aligns with previous research identifying uncertainty as the domain with the highest presence and severity scores [10, 15-18], and indicates the importance of measuring uncertainty as a core component of disability experiences [57]. Consistent with UK data, physical symptoms and impairments were the most episodic experienced by women in this sample [10, 16], emphasising the importance of screening, measuring and addressing these health challenges. Chronic pain of joints (45%) and soft tissue (42%) were the most frequent concurrent health conditions, with mobility impairments, social participation challenges and limitations with life activities being the most common WHODAS activity limitations. Musculoskeletal pain is common during perimenopause [58], highlighting the importance of detailed clinical history and awareness of the broad impacts of menopause transition. Furthermore, this sample of women reported higher rates of ≥1 WHODAS limitations than samples comprising mostly men, and higher rates of four or more activity limitations [3]. Physical symptoms and impairments can adversely affect daily functioning and reduce health-related quality of life [59]. Despite HIV care cascade successes in the UK [60], addressing the broader health, functioning, and well-being needs of women living with HIV requires disability-inclusive systems, interventions and multi-disciplinary care teams capable of providing comprehensive rehabilitation.

### Implications for practice and research

The EDQ can be used clinically to describe disability experienced by women living with HIV, determine the impact of health challenges, improve communication between providers and patients, and evaluate the effect of interventions [10]. Clinicians and women living with HIV can use the EDQ to better understand which disability dimensions pose challenges and better direct personalised interventions and rehabilitation. The EDQ is also unique in its ability to measure and describe the episodic nature of health challenges experienced over time [10, 61]. Our findings indicate women living with HIV in the UK are experiencing and self-reporting high levels of disability and multimorbidity. This warrants the inclusion of disability as a core outcome within national HIV surveillance, such as the UK Health Security Agency (UKHSA) annual reporting [1] and the Positive Voices survey [41], using validated tools to estimate prevalence and distribution of disability. Given that disability is associated with poorer health outcomes, fewer economic opportunities, and increased risk of poverty [62], the availability of valid and reliable disability data is essential for informing national strategies and action plans. Such data are critical for identifying priority areas and policy targets, guiding service planning and resource allocation, evaluating whether a population’s health needs are met, and assessing effective intervention coverage. Moreover, disability data are required for monitoring progress against the United Nations (UN) Sustainable Development Goals (SDGs) and for fulfilling obligations under the Convention on the Rights of Persons with Disabilities (CRPD) [63-66].

### Strengths and Limitations

We recruited ethnically diverse women living and ageing with HIV across the UK who had participated in the PRIME study, whereby the majority of participants were peri- or post-menopausal. Menopausal experiences of women in the UK are multifaceted, spanning physical, emotional, and social dimensions of health [67], and our results bring attention to the disability experiences of women living with HIV during the menopause. Our results may not be generalisable to all women living with HIV or transferable to low or middle-income countries. This assessment of EDQ properties involved electronic administration, and results may not be transferable to other modes of administration, such as paper-based administration. Women represent over a third of people receiving HIV care in the UK in 2024, with 48% aged 50 years and over [1]. Consequently, these data are representative of a large and growing population of women living with HIV in the UK, addressing chronic underrepresentation of women, racially minoritised people and older people in HIV research [22, 68].

## CONCLUSIONS

The EDQ possesses internal consistency, test-retest reliability, and construct validity with varied precision among women living with HIV. It can support research, clinical practice, and national policy to measure, describe and report disability, evaluate interventions, and identify priority areas for healthcare planning and allocation of services for women living with HIV in the UK.

## Data Availability

All relevant data are within the manuscript and its Supporting Information files.

## Acknowledgements

We express our deepest gratitude to all the women living with HIV who participated in this research, sharing their experiences and providing valuable insights into various aspects of their health and functioning. We also thank the research team, including co-investigators and collaborators from ten NHS outpatient HIV clinical settings recruiting participants, and the Sophie Forum for their dedicated efforts in providing patient and public involvement, for this evaluation of disability among PRIME study participants, the UK’s largest sample of women living with HIV.

